# Lesion network localization of functional and somatic symptoms

**DOI:** 10.1101/2025.03.06.25323494

**Authors:** Beatrice A. Milano, Stephan T. Palm, Matthew J. Burke, William Drew, Isaiah Kletenik, Andrew R. Pines, Maurizio Corbetta, Jordan H. Grafman, Jonathan Downar, Shan H. Siddiqi

## Abstract

Functional and somatic symptoms with no detectable structural abnormalities are a common cause of disability. These symptoms are widely believed to have neuropsychiatric origins, and thus may respond to network-targeted brain stimulation. To derive a network-based target, we studied functional and somatic disability after focal brain lesions. Using a normative human connectome database (n=1000), we mapped the circuitry functionally connected to lesions that selectively influence such symptoms in two datasets. First, in ischemic stroke (n=101), we mapped a network causally associated with self-reported functional disability, independent of individual measures of disability. In an independent sample with penetrating head trauma (n=181), lesions connected to our network were associated with greater somatic concern (p=0.001).

Across both datasets, functional and somatic symptoms were most associated with lesions connected to the orbitofrontal cortex (p_FWE_<0.01) and dorsal anterior cingulate, which we propose as potential brain stimulation targets.

## Introduction

Functional and somatic symptoms encompass a wide spectrum of reported somatoform concerns across medicine^1,2^. These symptoms can cause marked disability^3,4^, but often cannot be fully explained by structural, neurological, or other identifiable medical findings. Nevertheless, they occur across multiple disorders, such as functional neurological disorders (FND)^5–7^, chronic fatigue syndrome^8–10^, fibromyalgia^11^, irritable bowel syndrome^12^, and other chronic pain disorders. The debilitating nature of these disorders naturally entails extensive diagnostic testing and therapeutic attempts, which consequently leads to worldwide economic burden in healthcare ^13–15^.

Despite such ubiquity, FND and its related symptoms remain challenging to define and study mechanistically. Correlative neuroimaging studies have implicated multiple cognitive-behavioral brain networks in FND^14^. However, most of these findings were derived from correlative analysis, so it remains unclear if they are causally implicated in generating such clinical pictures. Some correlative findings may be causal, but they could also be compensatory, epiphenomenal, or reverse causal. Intervening on a compensatory process could be counterproductive, so causal substrates remain better therapeutic targets^16,17^.

Lesion studies can be used to identify causal neuroanatomical substrates as potential targets for focal brain stimulation^18^ . For instance, lesion-derived networks have been shown to yield better therapeutic brain stimulation targets for cognitive symptoms of Parkinson’s disease^19^, epilepsy^20^ , tics^21^, addiction^22^, depression^23^, and other disorders. Here, we studied patterns of brain lesions (n=282) associated with functional and somatic symptoms, as defined by self-assessed disrupted overall health, independent of specific limitations. We hypothesized that distinct brain connectivity patterns would be associated with more functional and somatic symptoms.

## Results

### Dataset characteristics

Baseline clinical characteristics and lesion locations are summarized in Figure 1 for both Dataset 1^24^ (Figure 1a-b) and Dataset 2^25^ (Figure 1c-d).

**Fig. 1.**
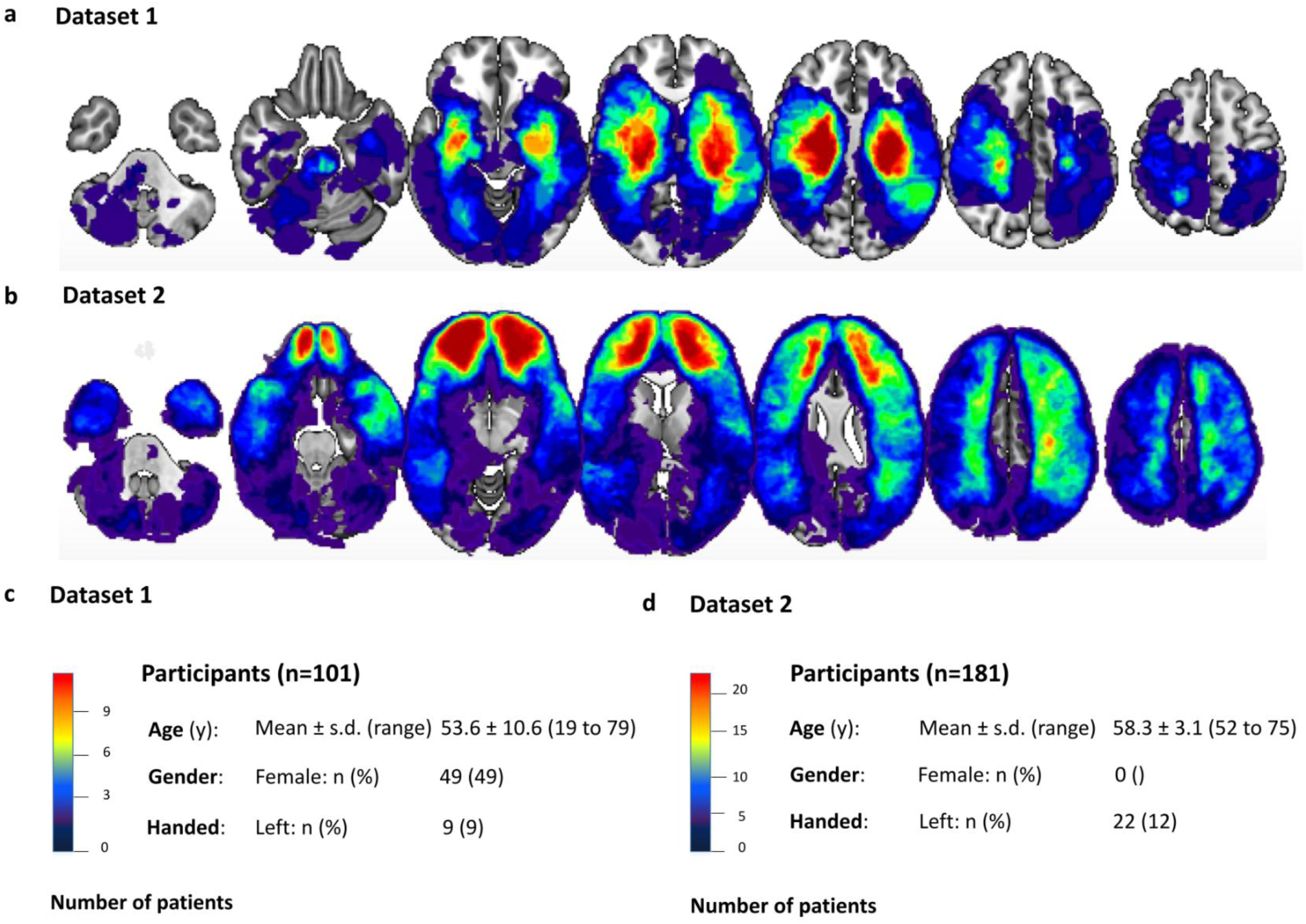
Distribution of the lesions and demographics in Dataset 1 and Dataset 2. a, Distribution of the lesions due to stroke across the brain in Dataset 1 b, Distribution of the lesions due to penetrating head trauma across the brain in Dataset 2 c, Demographics of Dataset 1 d, Demographics of Dataset 2

Dataset 1 included 101 patients who presented to an acute care hospital with ischemic stroke and no other neurological, psychiatric or medical conditions (i.e. dementia, depression, schizophrenia)^24^. Dataset 2 included 181 patients who sustained penetrating head trauma during the Vietnam War, and were evaluated over 15-20 years afterwards.

### Lesion modifying self-assessed functional disability are connected to a common brain network

101 participants (Dataset 1) with ischemic stroke lesions (Fig. 2a) completed the Short Form 36 Health Survey (SF-36)^26^ 3 months after the stroke. Stroke-induced change in functional symptoms was estimated using item SF-02, which asks: "*Compared to one year ago, how would you rate your current general health?*".

**Fig. 2.**
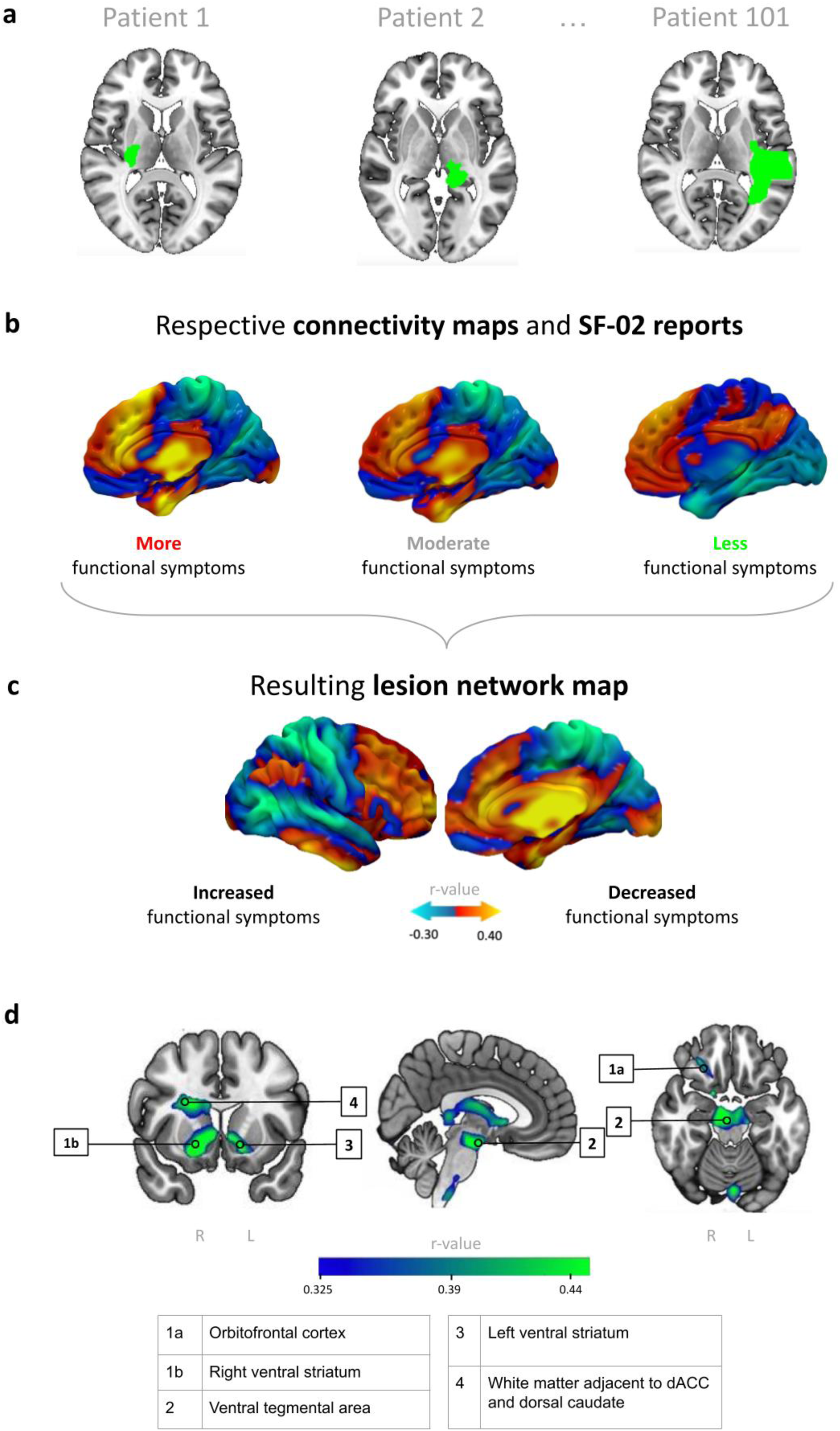
Research protocol: lesion network mapping of functional symptoms. a, Stroke lesions from Dataset 1 were segmented and warped to a standard brain atlas (n = 101 participants). Here we show three representative lesions. b, Voxel-based Pearson’s correlation: lesion connectivity was estimated using a normative connectome database (n = 1000). Normative connectivity maps of the three representative lesions are shown here. Computation of the respective connectivity maps (n=101) and data points are indicating quantified changes (SF-02 scores) in functional symptoms. Right hemisphere shown. c, Each lesion’s connectivity profile was compared with each patient’s functional symptoms score, yielding a network. Right hemisphere shown. d, Cluster-thresholded peaks of the LNM from Dataset 1 (detection threshold p < 0.001, cluster extent > 320 mm^3^). Full details can be retrieved in Table S3.

This item was chosen because it is the only question on the SF-36 that asks participants to compare current perceived overall health with that prior to the stroke event, with responses recorded on a 5-point Likert scale.

To map the network associated with lesion-induced changes in functional and somatic symptoms, we estimated the whole-brain connectivity of each lesion location using a human connectome database of healthy controls (n = 1000) (Fig. 2b).

This resulted in 101 connectivity profiles (t-maps) that were compared with changes to SF-02 score. At each voxel, we computed a partial Pearson correlation between lesion connectivity and SF-02 score, controlling for lesion size and the remaining items on the survey focused on quantifying specific limitations (i.e. physical, social, and emotional factors). These specific limitations were quantified using items such as "*To what extent has pain impeded your usual work activities?*", and "*Does your health condition constrain you in the activity of walking the distance of one block?*". The full list of SF-36 items is presented in Table S1. This analysis yielded a network map depicting the connectivity of brain lesions that was more likely to selectively and specifically modify self-assessed overall functioning (Fig. 2c). For convenience, we heretofore refer to this map as the functional symptoms network.

Of note, this analysis excluded item 1, items 13-16, and items 33-36, as these items partly focused on assessing overall health rather than specific dimensions, and could thus be highly collinear with the primary outcome. To ensure that the result was not biased by this methodological choice, we also repeated the analysis after including all items on the survey as covariates, which yielded a similar network (spatial r = 0.97).

We then identified cluster-thresholded peaks of the LNM from Dataset 1 with a detection threshold of p < 0.001 and a minimum cluster extent of 320 mm^3^ (Fig. 2d).

### External validation

Next, we validated our network in the independent cohort of patients from the Vietnam Head Injury Study (VHIS)^25^. 181 participants completed a head CT scan and the clinician-rated Neurobehavioral Rating Scale (NRS)^27^, which assesses the presence and chronicity of 27 different symptoms that commonly occur after brain injury. The NRS was completed after the rater had interacted with the participant for five full days of deep phenotyping. A full list of NRS questions is presented in Table S2.

Each lesion (Fig. 3a) was again mapped to its underlying connectivity profile (Fig. 3b), which in turn was compared with our functional symptoms network from Dataset 1 (Fig. 3c) using spatial correlation to estimate each lesion’s connectivity to such a network. Lesions with greater connectivity to our network were associated with greater somatic concern, as measured by item NRS-02 (r=0.23, p=0.0016) (Fig. 3d).

**Fig. 3.**
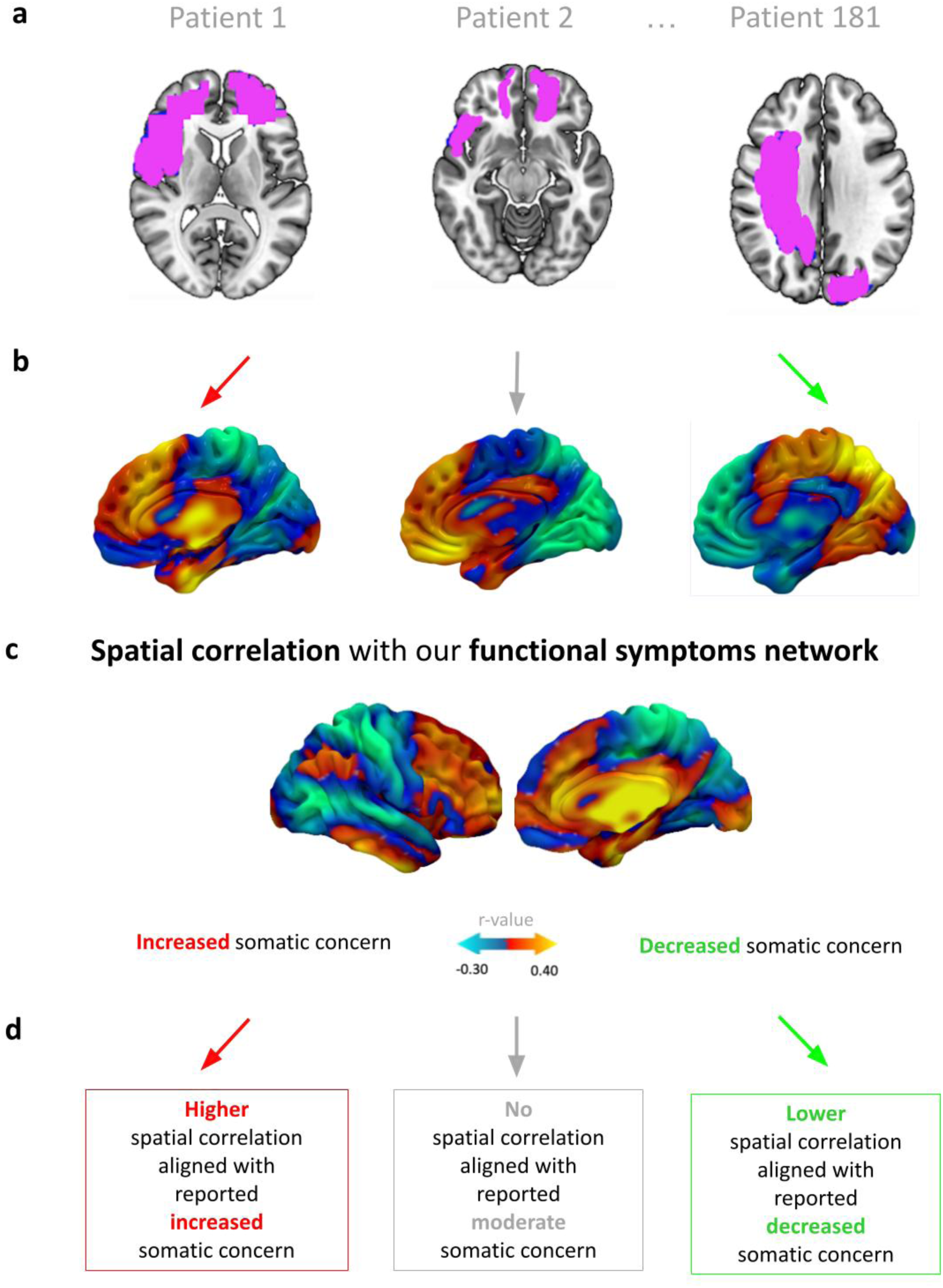
External validation of the functional symptoms network. a, Penetrating brain lesions from an independent dataset were automatically segmented and warped to a standard brain atlas (n = 181 participants). Here we show three representative lesions of, respectively, high, moderate, and absent somatic concern. b, Lesion connectivity was estimated using a normative connectome database (n = 1000). Normative connectivity maps of the three representative lesions are shown here.Right hemisphere shown. c, Each lesion’s connectivity profile was compared with our functional symptoms network (Fig.2c) using spatial correlation. Right hemisphere shown. d, Results of spatial correlation were compared with the Neurobehavioral Rating scale (NRS) using Spearman correlation.

The lesion network mapping procedure was also repeated for Dataset 2 (Fig. 4a). For convenience, we will heretofore refer to the resulting map as the somatic symptoms network. The resulting somatic symptoms network from Dataset 2 was highly similar to the functional symptoms network generated from Dataset 1 using spatial correlation (spatial r = 0.5853, p = 0.04). We identified cluster-thresholded peaks of the LNM from Dataset 2 using the same methods as Dataset 1 (Fig. 4a).

**Fig. 4.**
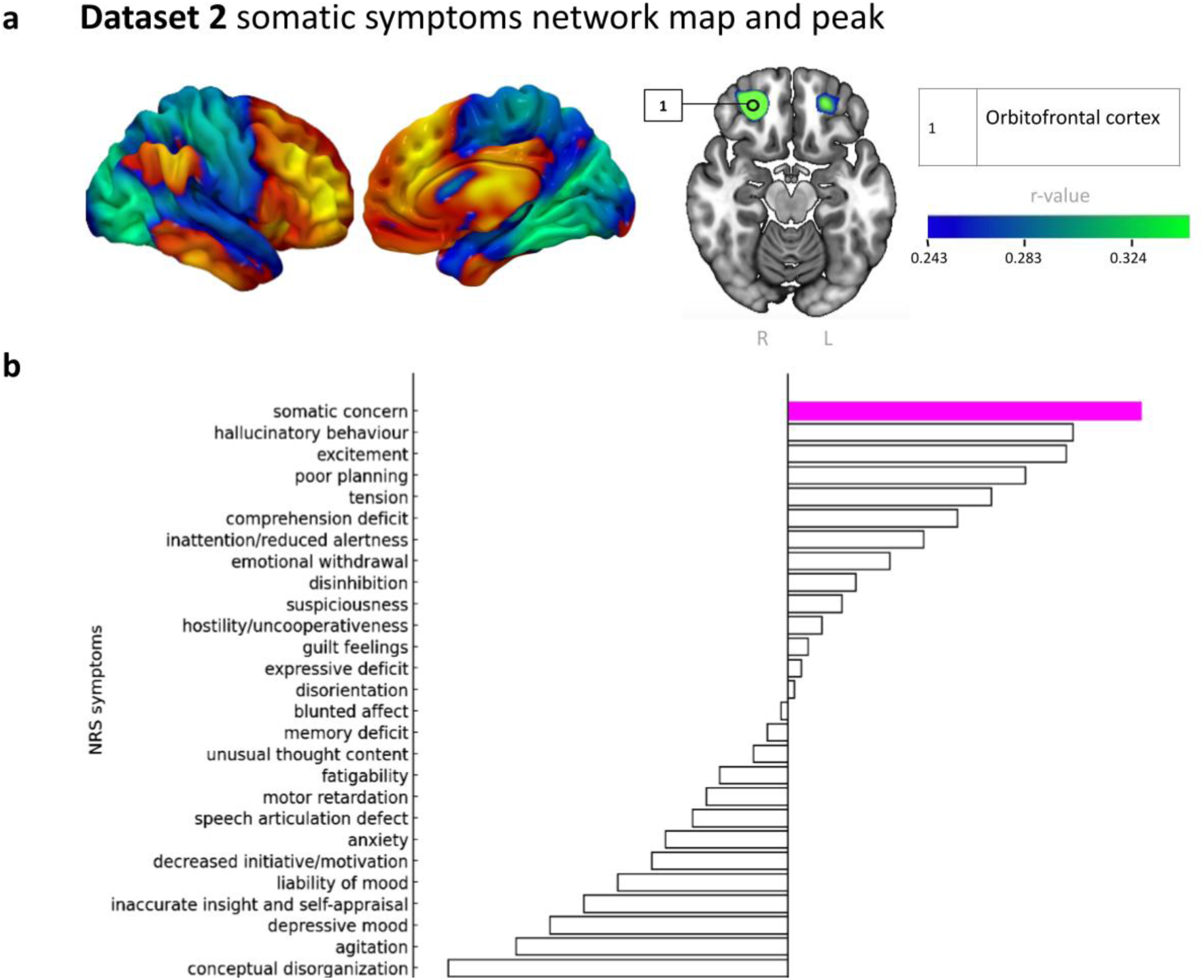
LNM from Dataset 2 and its predictive capacity. a, LNM from Dataset 2 (right hemipshere) and its cluster-thresholded peak (detection threshold p < 0.001, cluster extent > 320 mm^3^). More details can be found in the Supplementals (Table S3) b, The functional symptoms network from Dataset 1 showed higher spatial correlation with the somatic concern network from Dataset 2 than with any of the other NRS item-level networks from Dataset 2.

To test specificity, we repeated the lesion network mapping analysis for all of the remaining NRS items. The functional symptoms network map from Dataset 1 showed greater spatial correlation with the somatic symptoms network from Dataset 2 than any of the control maps from Dataset 2 (generated with all NRS items, except for NRS-02). In other words, somatic concern was the symptom that was most associated with lesion connectivity to the functional symptoms network (Fig. 4b).

### Combining both datasets and identifying potential targets for brain stimulation

As the functional symptoms network map from Dataset 1 (Fig. 5a) and the somatic symptoms network map from Dataset 2 (Fig. 5b) were significantly similar, we combined them into a weighted mean map (Fig. 5c) to generate an overall network associated with somatoform symptoms. For convenience, we refer to this as the functional and somatic symptoms network. Cluster extent thresholding (detection p < 0.001, cluster size > 320 mm3) revealed significant clusters in the lateral orbitofrontal cortex, ventral tegmental area, ventral striatum, and anterior limb of the internal capsule. All significant cluster locations from Dataset 1, Dataset 2 and combined Datasets are listed in the Supplement (Table S3).

**Fig. 5.**
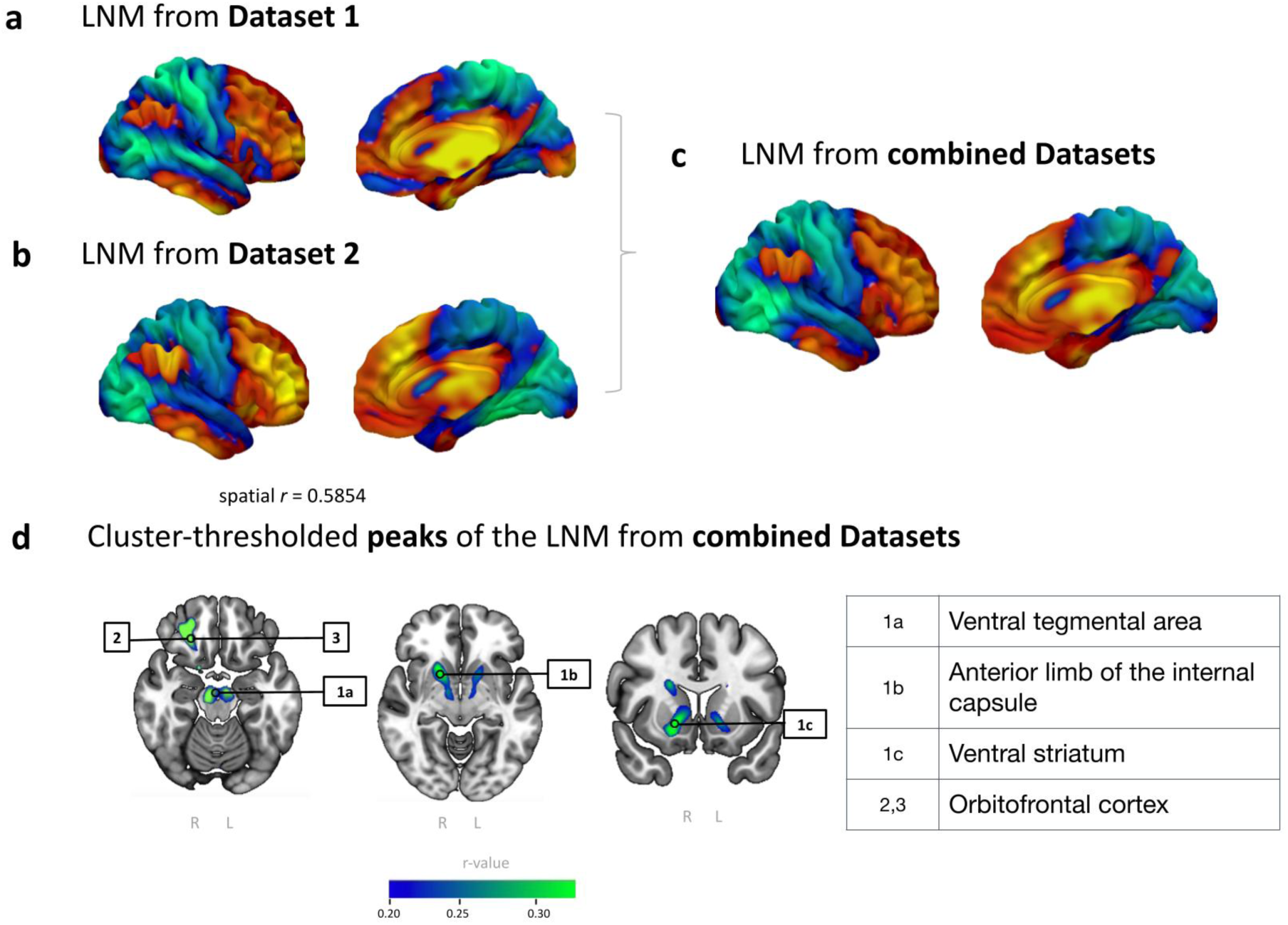
Our three LNMs. a, LNM from Dataset 1 (Right hemisphere shown) b, LNM from Dataset 2 (Right hemisphere shown) c, Combined and final LNM (Right hemisphere shown) d, Cluster-thresholded peaks of the LNM from combined Datasets (detection threshold p < 0.001, cluster extent > 320 mm^3^).

Because cluster extent thresholding may be excessively liberal^28^, we also used a more conservative permutation-based voxel-level multiple comparisons correction using the Westfall-Young method^29^ as implemented by Winkler et al.^30^. Of the clusters reported in Fig. 4d, the orbitofrontal cortex survived whole-brain voxel-level correction (p_FWE_ < 0.01).

Using a precomputed whole-brain connectome database, we then spatially correlated the functional and somatic symptoms network map with the whole-brain connectivity map of each voxel in the brain, as defined by the same normative connectome database. This results in a targeting atlas of potential brain stimulation targets based on how well their connectivity profile represents our network. The peak targets were in the pre-supplementary motor area (pre-SMA)/dACC [MNI 0, 32, 38], and OFC [MNI -28, 54, -10].

To determine the extent to which these TMS targets would overlap with the electrical field generated by a magnetic coil, we simulated TMS electric fields using SimNIBS^31^ . We employed a MagVenture B65 (MagVenture, Farum, Denmark) coil in MNI space using FpZ (dACC) and Fp1 (OFC) electrode placements on the 10-20 EEG system, sufficiently superficial to serve both as TMS targets (Fig. 6).

**Fig. 6.**
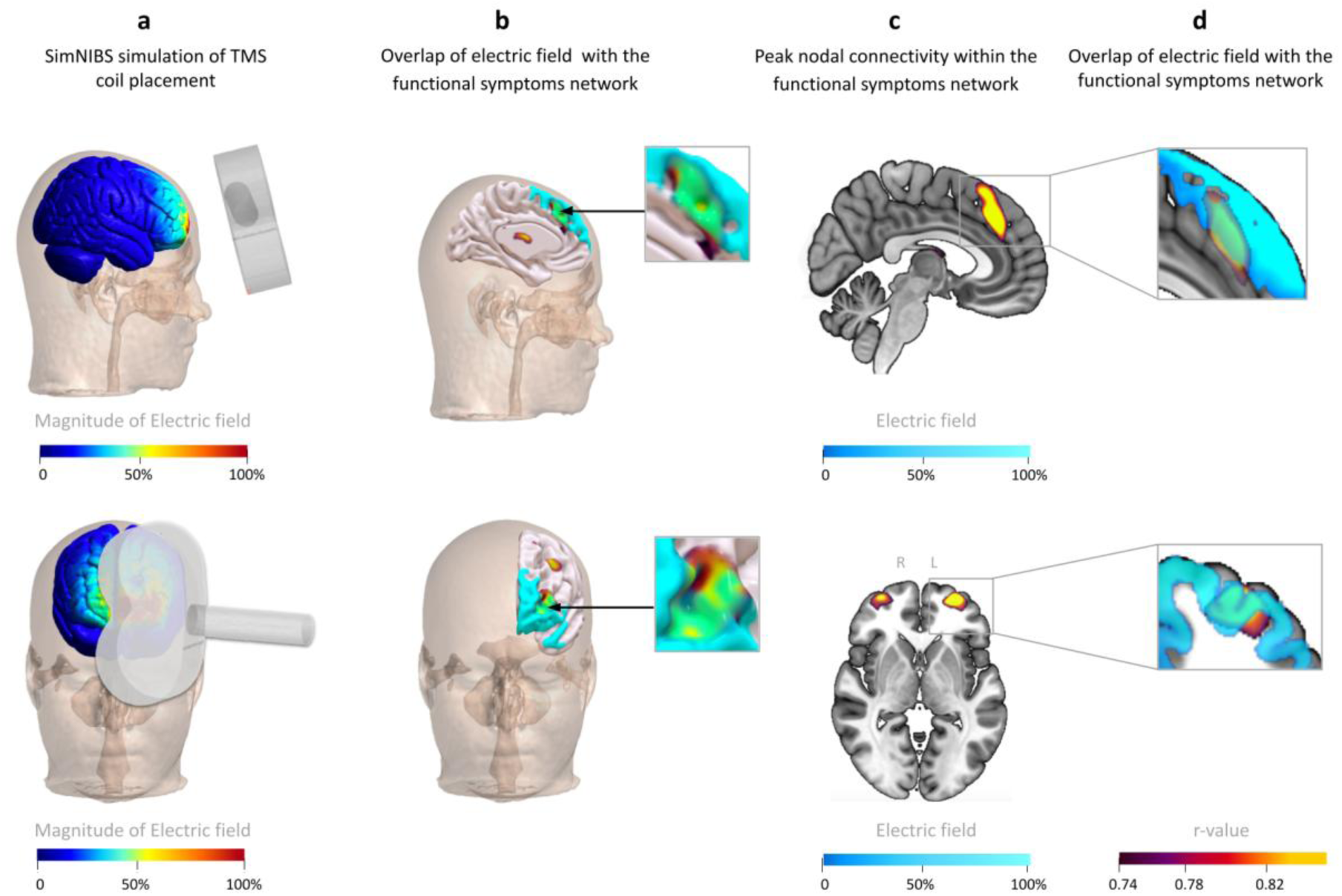
Potential therapeutic TMS targets identified by correlation with each voxel’s intrinsic functional connectivity. a, SimNIBS simulation of TMS coil placement, in FpZ (dACC) and Fp1 (OFC). b, Identification of voxels with the strongest correlation between intrinsic resting-state BOLD signal and BOLD signal specific to lesions that cause functional symptoms c, A simulated electric field measure with its two centers overlapping peak nodal connectivity within the network, indicating that some peaks are sufficiently superficial to be targeted with transcranial magnetic stimulation (TMS). d, Magnetic field generated by the TMS coil in the positions overlapping peak nodal connectivity within the network, identical to panel b, shown on a 3D model.

### Additional control analyses

Lesion network mapping (LNM) has also been used to localize a network for anosognosia^32^, or unawareness of one’s disability. It is plausible that our network may have reflected the absence of anosognosia. To explore this alternative explanation, we compared the two networks. Our functional and somatic symptoms network showed no spatial correlation with the anosognosia network (spatial r = -0.05, p= 0.93).

Of note, SF-36 was used as it contains metrics of both overall disability, but also of specific impairments, facilitating the relevant comparisons. However, it may not be the best metric of objective disability in functional somatic syndromes^33^. To address this, we repeated the analysis in Dataset 1 after controlling for other measures of objective disability. The network remained unchanged when controlling for NIH Stroke Scale^34^ (spatial r > 0.99), Functional Independence Measure^35^ (spatial r = 0.97), Functional Assessment Measure^36^ (spatial r = 0.99), Reintegration to Normal Living Index^37^ (spatial r = 0.95), Sickness Impact Profile^38^ (spatial r = 0.98) domains, and Geriatric Depression Scale ^39^ (spatial r = 0.99).

## Discussion

The subjective nature of functional and somatic symptoms has historically posed challenges across multiple clinical disciplines, raising debates about the relative contributions of neuropsychiatric and somatic components^40^. To our knowledge, this is the first study that uses causal sources of information to map functional and somatic symptoms to a reproducible brain network, independent of specific measures of physical disability, arguing in favor of a strong neuropsychiatric component.

This is consistent with emerging neuroimaging studies challenging the notion ^41,42^ that “functional” and/or “somatic” symptoms are defined by the absence of an anatomical (potentially measurable) substrate^14^. Furthermore, this also raises the possibility of treating FND and other somatoform disorders with network-based interventions such as TMS.

Although partially built on post-stroke functional connectivity data (Dataset 1), the functional symptoms network did not include areas traditionally associated with post-stroke disability, such as motor, sensory, language, or visual processing areas^24^.

Furthermore, the results survived correction for measures of general stroke severity, including lesion volume and clinical outcome measures such as NIH Stroke Scale. Thus, it appears unlikely that the functional symptoms network is driven by strokes that commonly cause marked physical disability, such as those affecting the middle cerebral artery.

Instead, the network clearly encompassed the VTA, ventral striatum (i.e. nucleus accumbens), ALIC, dACC and OFC. These regions are known to be integral for reward^43^, decision-making^44^, and learning^45^ across extensive human and animal studies. This suggests that functional and somatic symptoms may involve reward-related processes, although future research may investigate this further by specifically measuring reward-based learning and reward-motivated behavior.

Perhaps unsurprisingly, this anatomical distribution overlaps substantially with brain regions that have been extensively implicated in a range of psychiatric disorders. In particular, this also fits with the prevailing conceptualization of FND as a cognitive-behavioral syndrome rather than peripheral nerve pathology^7^. Major depressive disorder (MDD) has been associated with dysfunction of dopaminergic neurons in VTA, decreased dopamine concentration and metabolic rate in NAc/mPFC brain regions^46^, and dysregulated functional connectivity between dorsal limbic and neocortical areas^47^.

Obsessive-compulsive disorder (OCD) classically affects dACC, ALIC, and ventral striatum, all three of which have been used as neurosurgical treatment targets, respectively, with cingulotomy^48^, anterior capsulotomy^49^, and VC/VS DBS^50^. Similarly, addiction is associated with impaired dopamine signaling in the brain’s reward system, and, as the disorder progresses, decreased activity in OFC and ACC is often detected^51^.

The anatomical distribution of our network, particularly in dACC and lateral OFC, also partly overlaps with circuitry that has previously been implicated in pain^52^ and in placebo effect^53,54^, both of which share many similarities with FND. Chronic pain has also been framed as maladaptive integration of somatosensory and affective components^20^. These observations may suggest that pain-related and reward-related brain networks interact to dynamically tune the emotional salience ascribed to pain^55^. This interpretation is further supported by subsequent in-human data suggesting that OFC may integrate pain expectation and context-dependent predictions, influencing subjective pain evaluation^56^. Correlative functional MRI studies have similarly implicated reward-related regions, finding reduced connectivity between nucleus accumbens (NAc) and medial prefrontal cortex (mPFC)^57^ in chronic pain patients, and subsequent decreased VTA activity, which may induce motivational deficits and anhedonia^46^. Our network localization builds on these findings by providing causal directionality, suggesting that modulating the tendency to attend to pain may be a plausible approach with brain stimulation^18^.

Strengths of our study include permutation-based statistical assessments and robustness across two independent datasets with different lesion etiologies. There are also several limitations. The outcome measures were different across the two datasets - SF-02 used a self-report measure of perceived health, while NRS-02 used a clinician-report measure of somatic symptoms. As neither question (SF-02 and NRS-02) was directly asking about FND, we inferred it based on related constructs. It is not clear if any of these patients actually developed FND, so the link between FND and our functional and somatic symptoms network remains indirect. . Furthermore, we did not have pre-lesion data, and although patients in Dataset 1 were asked to compare their current status to their pre-stroke status, this may be subject to recall bias. Our analysis thus assumes that pre-lesion symptoms are unrelated to lesion location; while this may be a reasonable assumption, it likely introduced noise in the analysis. These limitations would be expected to reduce the observed effect size, so future studies may benefit from formally assessing for FND diagnosis and measuring the symptoms specific to the patient’s presentation. We identified a common network despite these sources of heterogeneity, suggesting that the network may subserve a transdiagnostic construct that can manifest as either functional or somatoform symptoms. Finally, while we propose a potential targeting atlas for TMS-induced changes in functional and somatic symptoms, this atlas is intended as a foundation for hypothesis generation, since it was not tested on TMS data. Future studies should seek to determine whether stimulation of different parts of this network can selectively induce clinically meaningful changes in functional symptoms.

In conclusion, we derived and replicated a brain network functionally connected to lesions that causally modify functional symptoms or somatic concern. This network overlaps with brain regions involved in motivation, reward, and learning, and is accessible with TMS. Future studies may employ the empirically shown targets in clinical trials for disorders that cause somatoform disability.

## Data Availability

All data produced in the present study are available upon reasonable request to the authors

## Acknowledgments

We express our gratitude to the participants who volunteered for the studies analysed in this work.

The authors would like to thank all research participants, funding bodies, healthcare personnel and other researchers who made this work possible.

This work was supported by the following sources: B.A.M.: Sant’Anna Scuola Superiore Universitaria Pisa (SFS); S.H.S.: National Institute of Health (grant nos. R01MH136248 and K23MH121657). M.C.: R01 HD061117-05

The funders were not directly involved in the conceptualization, design, analysis, decision to publish or preparation of the manuscript.

## Contributions

The conception and design of the study were by J.D. and S.H.S. Data collection was by M.C. and J.H.G. Preprocessing and preparation of data for analysis were by S.T.P., B.A.M., and W.D. The original draft of the manuscript was written by B.A.M. and S.H.S. Contributions to subsections and revisions of the final version of the manuscript were made by A.R.P., I.K., J.D., J.H.G.,M.J.B., S.T.P. and Simon Kwon. All illustrations were created by B.A.M. and S.H.S.

## Methods

### Datasets characteristics

#### Dataset 1

Stroke subjects (n=6260) were prospectively recruited from the stroke service at Barnes-Jewish Hospital (BJH) in St. Louis, Missouri, USA, with the help of the Washington University Cognitive Rehabilitation Research Group. After applying exclusion criteria (i.e., no history of stroke, no multifocal stroke, or no neurological/psychiatric history), 1209 met the inclusion criteria, and 172 were enrolled. Following further MRI-based exclusions and non-enrolment due to logistical reasons, the final study sample included 132 participants who completed the SF-36 questionnaire. Of these, our subset of interest concerned 101 individuals that responded to the SF-36 questionnaire^25^ at 3 months, enabling us to compare the current functional symptomatology to the patient’s recollection prior to the stroke.

Notably, neuropsychiatric and other medical conditions were criteria for exclusion. Stroke areas involved only middle and posterior cerebral artery territories.

Study design of data collection is described in its entirety in Corbetta et al. and supplemental materials^24^.

### Dataset 2

Dataset 2 originated from the Vietnam Head Injury Study (VHIS) registry, which includes 1221 American veterans who experienced brain injuries, primarily from penetrating head wounds during the Vietnam War. The registry, maintained by W.F. Caveness, contains data on veterans who sustained these injuries between 1967 and 1970, with detailed records completed by the attending neurosurgeon and follow-up information from the military and Veterans Administration (VA). Approximately 15 years later, the 1118 veterans still alive were invited to participate in an extensive follow-up clinical study. Recruitment, transportation, and the study itself were coordinated by the VA, the Armed Services, and the American Red Cross.

Our subset of interest included 181 participants, namely, those who responded to the NRS questionnaire were selected for our study.

Study design of data collection is described in its entirety in Grafman et al. and supplemental materials^25^.

#### Outcome measures: SF-36 and NRS

The Short Form 36-Item Health Survey (SF-36)^26^, devised by the RAND Corporation, is a questionnaire that evaluates health-related quality of life. The items on the survey are listed in Supplemental Table 1. Comprising a set of 36 items, it consists of eight health domains. These include physical functioning, bodily pain, emotional well-being, and social interactions.

The SF-36 employs a two-step scoring methodology, wherein responses are quantified to yield domain-specific scores. These scores facilitate cross-sectional and longitudinal comparisons.

Among its items, question SF-02 was chosen because it asks respondents to compare their overall health between the current time and one year earlier. By studying this survey 3 months after a stroke, participants are implicitly comparing their health before and after the stroke. A higher score on SF-02 indicates a greater perceived degree of functional symptoms.

We controlled for physical limitations in daily activities (SF-03 to SF-12) to ensure that the relationship was not driven by specific daily physical challenges. Similarly, we also controlled for the impact of emotional health/well-being, pain, and social functioning (SF-17 to SF-32). SF-01 and SF-33 to SF-36 were not included as covariates because they also ask about general health, which would be redundant with SF-02.

The Neurobehavioral Rating Scale (NRS)^27^, which studies a range of cognitive and emotional processes, was completed by a clinician after interacting with the participant for five days of deep phenotyping. Specifically, a higher score on NRS-02 indicates greater somatic concern. Other cognitive domains include depression, anxiety, attention, memory, executive functions, social interactivity, and language abilities.

### Lesion network mapping

#### Lesion analysis and functional connectivity mapping

As in our group’s prior work^20,22^, we utilized a normative human connectome database comprising resting-state fMRI scans from 1000 individuals as a reference to examine whole-brain connectivity associated with specific lesion sites. The analysis was conducted using MATLAB 2023a (MathWorks, Inc., Natick, Massachusetts, USA), employing custom-built scripts to compute the functional connectivity maps for the identified lesion areas. We processed lesion masks from Dataset 1, consisting of 101 individuals. Lesion masks were loaded, and the size of each lesion (i.e., the number of voxels with positive values) was calculated.

Time courses of voxels within the identified lesions were averaged, and these averages were correlated with the time courses of all other brain voxels. This step yielded individual connectivity maps for each lesion, which were then averaged across all subjects to produce a single functional connectivity map per lesion.

### Correlation with functional symptoms

Next, we estimated the correlation between the connectivity maps and the change in functional symptoms following a stroke, as quantified with SF-02 score. Using Pearson’s partial correlation, controlling for lesion size and remaining non-redundant SF-36 items, we calculated the correlation between the scores and the connectivity maps for each lesion.

This resulted in a whole-brain network specific for functional symptoms.

### Cross-dataset spatial correlation

To test for robustness, we generated a second network map following the same procedure for Dataset 2, which included 181 individuals with traumatic brain injury. Then we estimated Pearson’s correlation between the connectivity maps and the change in somatic concern, as quantified with NRS-02 score, controlling for lesion size and remaining NRS items. The two maps were Fisher z-transformed and then spatially correlated to assess their similarity.

To test this spatial correlation for significance, we used a permutation test, as in our prior work. We repeated the full analysis after randomly permuting each patient’s imaging against a different patient’s clinical outcomes within the same dataset. This yielded a distribution of spatial correlation values expected by chance, across 10000 iterations. A p-value was computed as the percentage of iterations in which the permuted spatial correlation exceeded the real spatial correlation. This non-parametric method is necessary because brain voxels are not independent of one another, and thus spatial correlations cannot be tested for significance using parametric methods.

### Testing specificity of the network

We repeated the lesion network mapping procedure for all NRS items. Pearson’s partial correlations were calculated between lesion connectivity maps and each NRS item, while controlling for others, generating specific lesion connectivity maps. Each NRS item was also mapped without controlling for others. We hypothesized that the functional symptoms map from Dataset 1 would better resemble the somatic concern map from Dataset 2 than any of the other symptom maps from Dataset 2.

### Combined network generation

We combined the connectivity maps from both Datasets by calculating a weighted average, based on sample sizes, to create a unified map. This combined network was then used for further analysis.

### Localization of significant clusters

To identify which voxels were significantly stronger than chance, we used the permutation-based Westfall-Young^29^ correction for multiple comparisons, as implemented by Winkler et al^30^. The overall network was recalculated after randomly permuting each patient’s clinical outcome with a different patient’s neuroimaging, and the peak t-value was recorded in 10000 random iterations. Voxels were considered significant if they were stronger than at least 95% of the peaks generated from this permutation procedure. Relevant brain regions were identified using the Harvard-Oxford cortical/subcortical structural atlas^58^ for the cerebrum.

### Identification of sites for stimulation

Using the same normative functional connectome database, we mapped the functional connectivity of every voxel, and computed its spatial correlation with the functional symptoms circuit. This yields a map of how similar each voxel’s connectivity profile is to the functional symptoms network, thus providing an atlas of brain regions that could potentially be targeted to maximize stimulation site connectivity to the network. To assess the extent to which targeting the dACC and OFC with TMS would align with the electrical field generated by a magnetic coil, we modeled the electrical field (E-field) in MNI space for a MagVenture B65 coil (MagVenture, Farum, Denmark) positioned at FpZ and Fp1, using SimNIBS^31^.

---

**Table S1.** 36-Item Short Form Survey Instrument (SF-36)

|  |  |  |  |  |  |  |
| --- | --- | --- | --- | --- | --- | --- |
| 1. In general, would you say your health is: | Excellent | Very good | Good | Fair | Poor |  |
|  | O | O | O | O | O |  |
| 2. Compared to one year ago, how would you rate your health in general now? | Much better | Much better | Much better | Much better | Much worse |  |
|  | O | O | O | O | O |  |
| 3. Does your health now limit you in vigorous activities, such as running, lifting heavy objects, participating in strenuous sports? | Yes, limited a lot | Yes, limited a little | No, not limited at all |  |  |  |
|  | O | O | O |  |  |  |
| 4. Does your health now limit you in vigorous activities, such as running, lifting heavy objects, participating in strenuous sports?<br>Does your health now limit you in moderate activities, such as moving a table, pushing a vacuum cleaner, bowling, or playing golf? | Yes, limited a lot | Yes, limited a little | No, not limited at all |  |  |  |
|  | O | O | O |  |  |  |
| 5. Does your health now limit you in lifting or carrying groceries? | Yes, limited a lot | Yes, limited a little | No, not limited at all |  |  |  |
|  | O | O | O |  |  |  |
| 6. Does your health now limit you in climbing several flights of stairs? | Yes, limited a lot | Yes, limited a little | No, Not limited at all |  |  |  |
|  | O | O | O |  |  |  |
| 7. Does your health now limit you in climbing one flight of stairs? | Yes, limited a lot | Yes, limited a little | No, not limited at all |  |  |  |
|  | O | O | O |  |  |  |
| 8. Does your health now limit you in bending, kneeling, or stooping? | Yes, limited a lot | Yes, limited a little | No, not limited at all |  |  |  |
|  | O | O | O |  |  |  |
| 9. Does your health now limit you in walking more than a mile? | Yes, limited a lot | Yes, limited a little | No, not limited at all |  |  |  |
|  | O | O | O |  |  |  |
| 10. Does your health now limit you in walking several blocks? | Yes, limited a lot | Yes, limited a little | No, not limited at all |  |  |  |
|  | O | O | O |  |  |  |
| 11 Does your health now limit you in walking one block? | Yes, limited a lot | Yes, limited a little | No, not limited at all |  |  |  |
|  | O | O | O |  |  |  |
| 12. Does your health now limit you in bathing or dressing yourself? | Yes, limited a lot | Yes, limited a little | No, not limited at all |  |  |  |
|  | O | O | O |  |  |  |
| 13. During the past 4 weeks, have you cut down the amount of time you spent on work or other activities as a result of your physical health? | Yes | No |  |  |  |  |
|  | O | O |  |  |  |  |
| 14. During the past 4 weeks, have you accomplished less than you would like as a result of your physical health? | Yes | No |  |  |  |  |
|  | O | O |  |  |  |  |
| 15. During the past 4 weeks, were you limited in the kind of work or other activities as a result of your physical health? | Yes | No |  |  |  |  |
|  | O | O |  |  |  |  |
| 16. During the past 4 weeks, have you had difficulty performing work or other activities (for example, it took extra effort)? | Yes | No |  |  |  |  |
|  | O | O |  |  |  |  |
| 17. During the past 4 weeks, have you had difficulty performing work or other activities (for example, it took extra effort)? | Yes | No |  |  |  |  |
|  | O | O |  |  |  |  |
| 18. During the past 4 weeks, have you accomplished less than you would like as a result of any emotional problems? | Yes | No |  |  |  |  |
|  | O | O |  |  |  |  |
| 19. During the past 4 weeks, have you not done work or other activities as carefully as usual as a result of any emotional problems?. | Yes | No |  |  |  |  |
|  | O | O |  |  |  |  |
| 20. During the past 4 weeks, to what extent has your physical health or emotional problems interfered with your normal social activities with family, friends, neighbors, or groups? | Not at all | Slightly | Moderately | Severe | Very severe |  |
|  | O | O | O | O | O |  |
| 21. How much bodily pain have you had during the past 4 weeks? | None | Very mild | Mild | Moderate | Severe | Very severe |
|  | O | O | O | O | O | O |
| 22. During the past 4 weeks, how much did pain interfere with your normal work (including both work outside the home and housework)? | Not at all | A little bit | Moderately | Quite a bit | Extremely |  |
|  | O | O | O | O | O |  |
| 23. During the past 4 weeks, how much of the time did you feel full of pep? | All of the time | Most of the time | A good bit of the time | Some of the time | A little bit of the time | None of the time |
|  | O | O | O | O | O | O |
| 24. During the past 4 weeks, have you been a very nervous person? | All of the time | Most of the time | A good bit of the time | Some of the time | A little bit of the time | None of the time |
|  | O | O | O | O | O | O |
| 25. During the past 4 weeks, have you felt so down in the dumps that nothing could cheer you up? | All of the time | Most of the time | A good bit of the time | Some of the time | A little bit of the time | None of the time |
|  | O | O | O | O | O | O |
| 26. During the past 4 weeks, have you felt calm and peaceful? | All of the time | Most of the time | A good bit of the time | Some of the time | A little bit of the time | None of the time |
|  | O | O | O | O | O | O |
| 27. During the past 4 weeks, did you have a lot of energy? | All of the time | Most of the time | A good bit of the time | Some of the time | A little bit of the time | None of the time |
|  | O | O | O | O | O | O |

**Table S1 | 36-Item Short Form Survey Instrument (SF-36)**
| 36-Item Short Form Survey Instrument (SF-36) |  |  |  |  |  |  |
| --- | --- | --- | --- | --- | --- | --- |
| 28. During the past 4 weeks, have you felt downhearted and blue? | All of the time | Most of the time | A good bit of the time | Some of the time | A little bit of the time | None of the time |
|  | ☐ | ☐ | ☐ | ☐ | ☐ | ☐ |
| 29. During the past 4 weeks, did you feel worn out? | All of the time | Most of the time | A good bit of the time | Some of the time | A little bit of the time | None of the time |
|  | ☐ | ☐ | ☐ | ☐ | ☐ | ☐ |
| 30. During the past 4 weeks, have you been a happy person? | All of the time | Most of the time | A good bit of the time | Some of the time | A little bit of the time | None of the time |
|  | ☐ | ☐ | ☐ | ☐ | ☐ | ☐ |
| 31. During the past 4 weeks, did you feel tired? | All of the time | Most of the time | A good bit of the time | Some of the time | A little bit of the time | None of the time |
|  | ☐ | ☐ | ☐ | ☐ | ☐ | ☐ |
| 32. During the past 4 weeks, how much of the time has your physical health or emotional problems interfered with your social activities (like visiting with friends, relatives, etc.)? | All of the time | Most of the time | Some of the time | A little bit of the time | None |  |
|  | ☐ | ☐ | ☐ | ☐ | ☐ |  |
| 33. Do you seem to get sick a little easier than other people? | Definitely true | Mostly true | Don't know | Mostly false | Definitely false |  |
|  | ☐ | ☐ | ☐ | ☐ | ☐ |  |
| 34. Are you as healthy as anybody you know? | Definitely true | Mostly true | Don't know | Mostly false | Definitely false |  |
|  | ☐ | ☐ | ☐ | ☐ | ☐ |  |
| 35. Do you expect your health to get worse? | Definitely true | Mostly true | Don't know | Mostly false | Definitely false |  |
|  | ☐ | ☐ | ☐ | ☐ | ☐ |  |
| 36. Is your health excellent? | Definitely true | Mostly true | Don't know | Mostly false | Definitely false |  |
|  | ☐ | ☐ | ☐ | ☐ | ☐ |  |

**Table S2.**
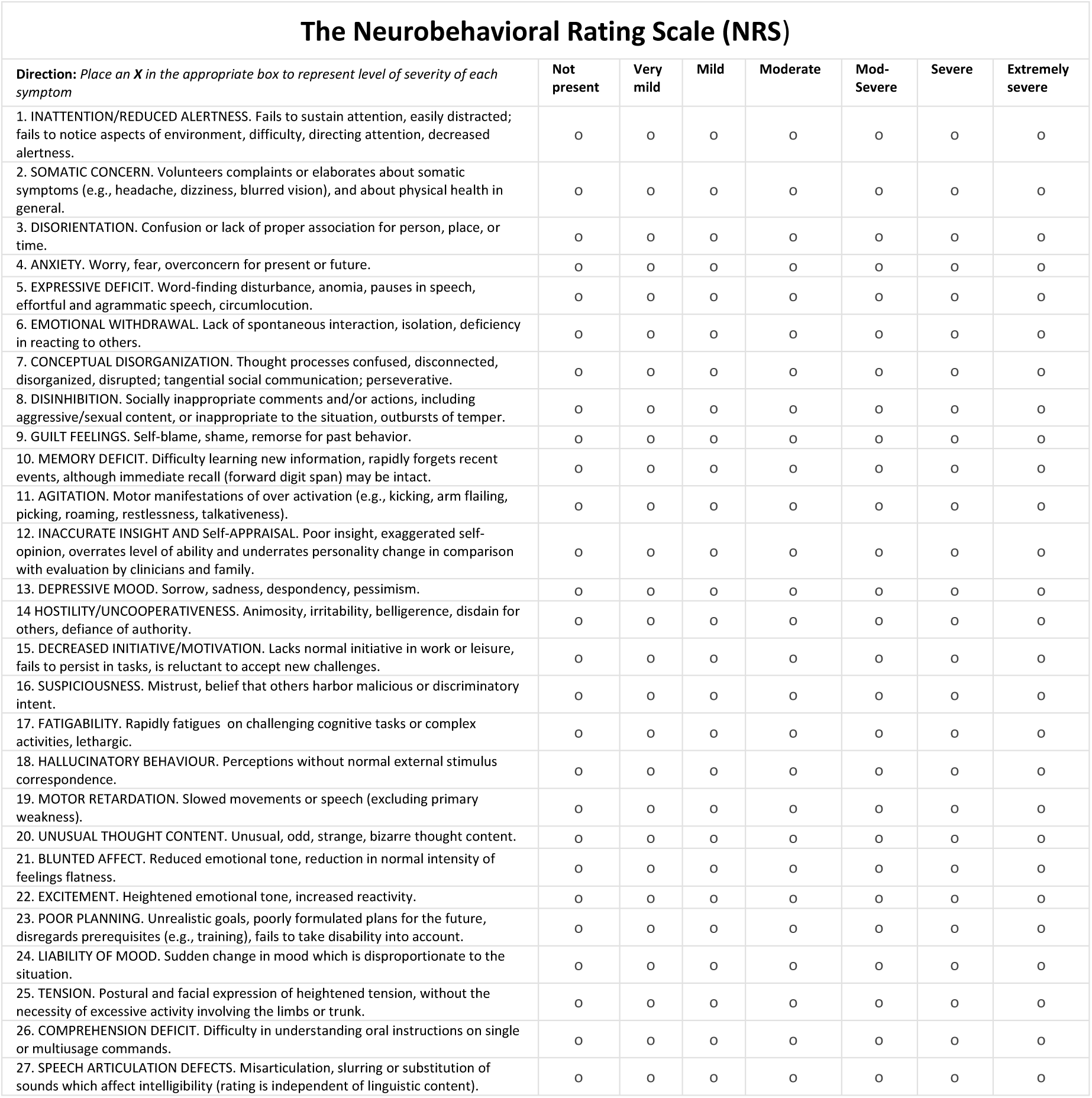
The Neurobehavioral Rating Scale (NRS)

**Table S3.** Cluster-thresholded peaks of the three network maps (detection threshold p < 0.001, cluster extent of 40 voxels = 320 mm^3^)

| Datasets | Cluster extent in voxels<br>($> 320 \text{ mm}^3$) | Anatomical areas included in each cluster | Peak MNI coordinate |
| --- | --- | --- | --- |
| 1 = functional symptoms after stroke<br>2 = somatic symptoms after TBI |  |  |  |
| 1 | 287 | OFC, nucleus accumbens, right ventral striatum | (14, 8, -8) |
| 1 | 60 | Left ventral striatum | (-12, 4, -8) |
| 1 | 49 | VTA | (8, -14, -18) |
| 1 | 40 | White matter adjacent to dACC and dorsal caudate | (20, 4, 26) |
| 2 | 141 | OFC | (24, 44, -20) |
| Combined | 2176 | VTA, ALIC, Ventral striatum | (6, 0, 6) |
| Combined | 374 | OFC | (24, 38, -22) |
| Combined | 173 | OFC | (-26, 38, -24) |

